# Towards Harmonizing Quantification of Dopamine Neuron Imaging Biomarkers in Parkinson’s Disease: The Centamine Scale

**DOI:** 10.1101/2025.08.19.25333801

**Authors:** Zhen Fan, Graham E. Searle, Gaia Rizzo, Justin Albani, Patrick A Cella, Robert A. Comley, Gregory Klein, Luca Passamonti, Cristian A. Salinas, Adam J Schwarz, Leonardo Iaccarino, Gilles Tamagnan, Jamie L. Eberling, Ken Marek, John P. Seibyl, Roger N. Gunn, The Parkinson’s Progression Markers Initiative

**Author notes:** Corresponding author: Roger N. Gunn 3 Portobello Terrace, North Worple Way, London, SW14 8AF Corresponding author’s.

## Abstract

**Objective:** Dopaminergic imaging is a key biomarker for both the investigation of biology of Parkinson’s Disease and related synucleinopathies and the evaluation of potential therapies in clinical trials. This work presents a harmonized approach for quantifying dopaminergic molecular imaging tracers, such as [¹²³I]ioflupane (DaTscan™) SPECT and [¹[F]AV133 PET, which assess dopaminergic neuronal loss. The proposed method aims to standardize regional outcome measures using a unified scale called Centamines.

**Methods:** The Centamines framework comprises three analysis levels. Level 1 defines the Centamine scale based on healthy subject data from [¹²³I]ioflupane SPECT (n=224). Level 2 uses Head-to-Head data between Tracer X and [¹²³I]ioflupane SPECT to map Tracer X onto the Centamine scale. Level 3 maps additional tracers using prior mappings. A Level 2 analysis was performed using [¹²³I]ioflupane SPECT and [¹[F]AV133 PET data (n=68) to convert [¹[F]AV133 PET into Centamines.

**Results:** Level 1 successfully established the Centamine scale using healthy [¹²³I]ioflupane SPECT scans. Level 2 revealed moderate-strong linear correlations (R² = 0.44–0.78) between [¹²³I]ioflupane SPECT and [¹[F]AV133 PET across five brain regions. Mapped Centamine values showed minimal differences between tracers, ranging from 1.5% (Post-Commissural Putamen) to 3% (Caudate).

**Interpretation:** The Centamine scale holds promise for the harmonized quantification of dopaminergic neuronal imaging markers. The Centamine strategy would enable and accelerate clinical trials in Parkinson’s Disease utilizing dopaminergic imaging outcomes.

## INTRODUCTION

Recently, data from the Parkinson Progression Marker initiative (PPMI) have demonstrated that the α-synuclein seed amplification assay (asyn SAA) coupled with dopamine transporter (DAT) imaging provide a biomarker driven definition for Parkinson’s Disease (PD) ^1^. This has led to the introduction of the concept of Neuronal Synuclein disease (NSD) encompassing PD, dementia with Lewy Bodies and REM Sleep Behavior Disorder (RBD) and the NSD-Integrated Staging System (NSD-ISS; ^2^). A similar proposal by the *SynNeurGe* group also supports a biologic classification for PD ^3^. A key goal for the biologic definition and staging is to establish biomarker strategies that inform and accelerate therapeutic trials at all stages of disease.

Dopaminergic imaging biomarkers are well established for PD. Among them, the DAT imaging agent [¹²³I]Ioflupane Single-photon emission computed tomography (SPECT) is the most widely used ^4–6^, with DAT PET imaging agents such as [¹[F]FPCIT ^7,8^ and [¹[F]PE2I ^9,10^ also routinely deployed in research studies. More recently, [¹[F]AV133, a PET biomarker targeting Vesicular Monoamine Transporter 2 (VMAT2; ^11–13^), has shown enhanced sensitivity for longitudinal monitoring of dopaminergic integrity ^14^.

In a related field, recent efforts focused on amyloid and tau imaging have developed a harmonized molecular imaging quantification approach. In 2015 the Centiloid quantification strategy was introduced ^15^ for the harmonized analysis of Amyloid PET tracers including [^11^C]PIB, [^18^F]Florbetapir, [^18^F]Flutemetamol and [^18^F]Florbetaben. This was the first time a universal quantification method had been developed for different molecular imaging agents targeting the same biological entity. It has been highly successful in improving the deployment and utility of amyloid imaging in Alzheimer’s disease (AD) clinical trials ^16^. By enabling use of multiple tracers, the method has facilitated the execution of large-scale trials while providing a standardized and easily interpretable scale for eligibility assessment and longitudinal data reporting ^17,18^.

Building on the success of Centiloids, the Critical Path for Alzheimer’s Disease (CPAD) has spearheaded the development of a similar approach for tau quantification, known as Centaurs^19^. This framework harmonizes quantification across established tau PET tracers, further enabling their use in clinical trials and enhancing their future clinical utility.

Given these advances, establishing a harmonized quantification approach for dopamine imaging agents would significantly enhance their application in PD clinical therapeutic trials, potentially improving the accuracy of cohort recruitment and assessment of treatment efficacy. This paper introduces the Centamine scale, detailing its framework and methodology, and presents data from [¹²³I]Ioflupane SPECT and [¹[F]AV133 PET to illustrate the approach.

## METHODS

### Data Sets

The data sets used in this work are part of the PPMI study (http://ppmi-info.org). All [¹²³I]Ioflupane SPECT and [^18^F]AV133 PET data were acquired according to the PPMI protocols ([¹²³I]Ioflupane SPECT: Dose: 111-185 MBq, Acquisition: 30min at 4hr post injection; [^18^F]AV133 PET: Dose: 185 MBq +/- 10%, Acquisition: 10min at 80min post injection.^20,21^ This analysis used data openly available from PPMI (Tier 1).

- Data Set 1 (Healthy [¹²³I]Ioflupane SPECT): 227 healthy individuals (Sex: 137 males/ 90 females, Age: 30-85 yrs, m: 62.0, sd: 11.8 yrs) were imaged with [¹²³I]Ioflupane SPECT.
- Data Set 2 (Head-to-Head [¹²³I]Ioflupane SPECT and [^18^F]AV133 PET); 68 individuals (51 Sporadic PD, 9 Hyposmia, 2 LRRK2, 2 Healthy Controls, 1 GBA, 1 PRKN, 1 RBD, 1 SWEDD; Sex: 43 males / 25 females, Age: 33-83 yrs, m: 64.0, sd: 9.1 yrs) were imaged with both [¹²³I]Ioflupane SPECT and [^18^F]AV133 PET. The interval between Head-to-Head scans was no more than 100 days and 45 subjects had multiple Head-to-Head acquisitions as part of a longitudinal assessment over a two-year period that led to a total of 162 paired scans.

An example of [¹²³I]Ioflupane SPECT and [^18^F]AV133 PET imaging data obtained in the same subjects for one healthy and three PD subjects are shown in Figure 1, highlighting the differences in spatial resolution provided by the two modalities.

**Figure 1.**
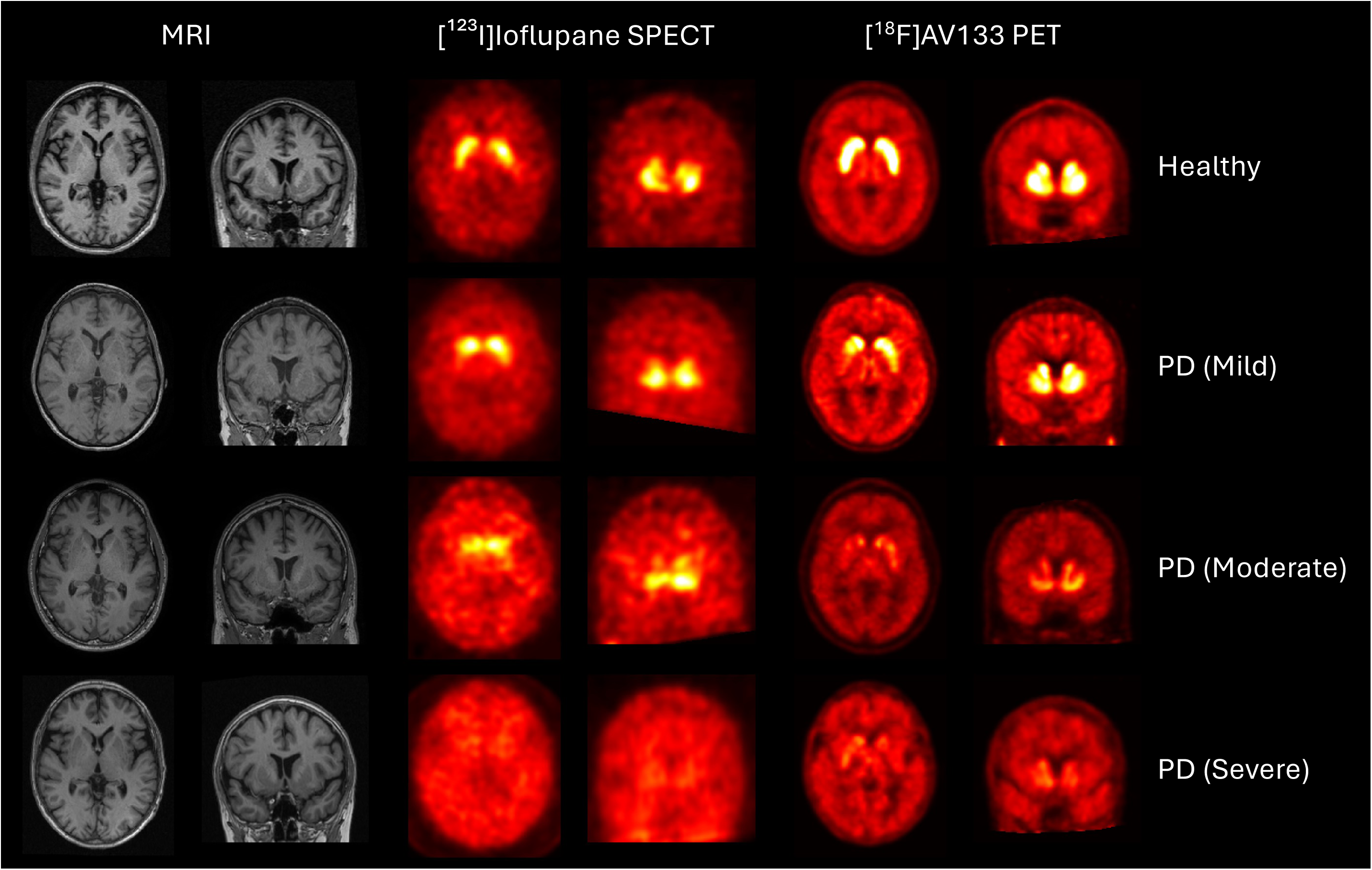
[¹²³I]Ioflupane SPECT (DAT), [^18^F]AV133 PET (VMAT2) and associated structural T1-MRI images for a healthy subject, mild, moderate and severe PD subject.

### Analysis Pipelines

All quantitative analyses were performed in MIAKAT^TM^ (Version 5.0)^22^. A standardised set of anatomical regions relevant to dopaminergic imaging defined in the CIC atlas ^23^ were used (Striatum, Putamen, Caudate, Pre-Commissural Putamen, Post-Commissural Putamen) and the reference region was chosen to be the cerebral white matter (CWM) as this has demonstrated increased performance over occipital reference region approaches ^14^. These target and reference regions are illustrated in Figure 2.

**Figure 2.**
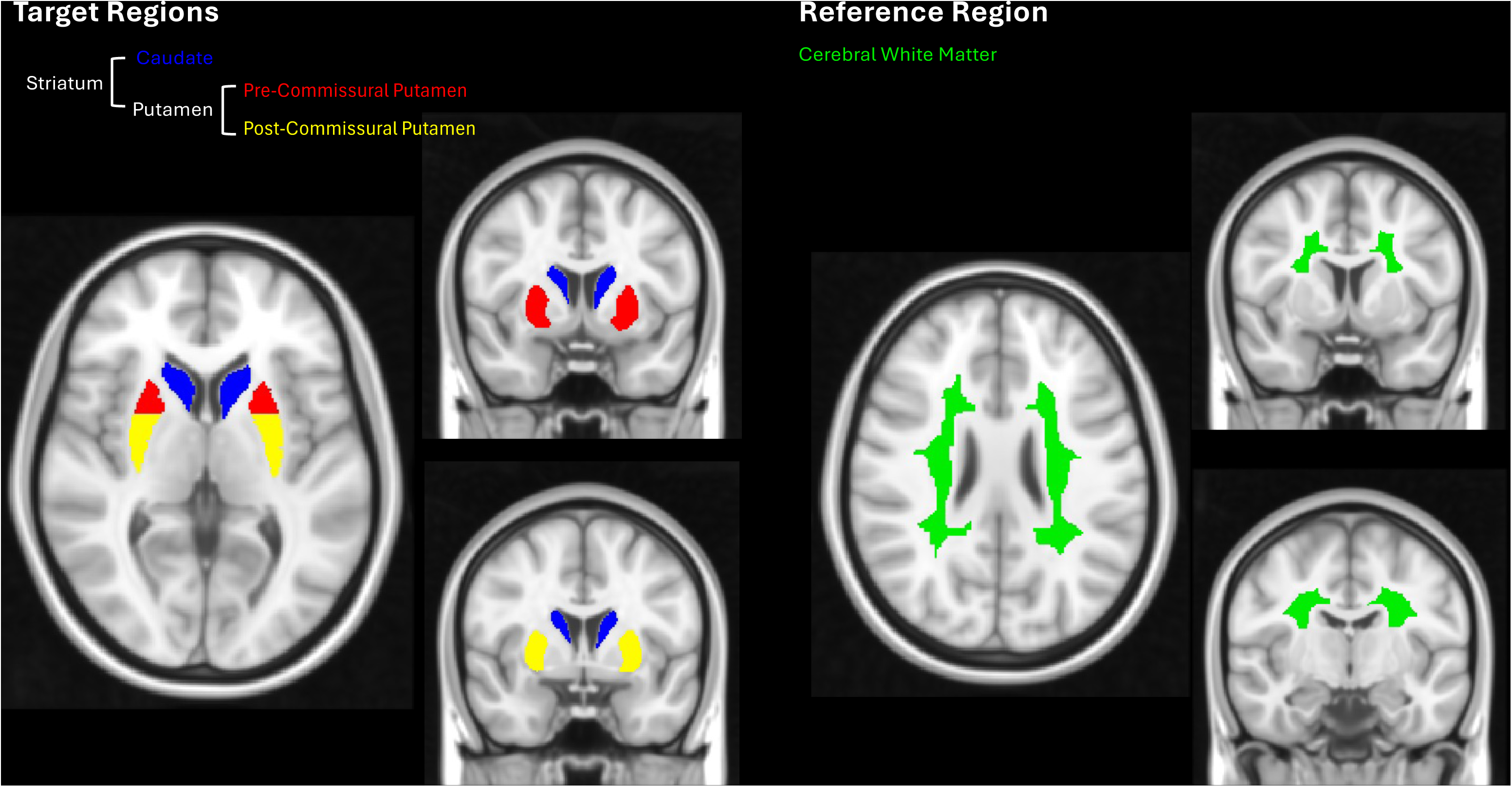
Target and Reference regions defined in MNI152 space.

### [¹²³I]Ioflupane SPECT Analysis Pipeline

SPECT imaging, such as [¹²³I]Ioflupane imaging, presents challenges in spatial registration and normalization due to the low resolution which can impact the accuracy of signal quantification. Given that most SPECT studies acquire [¹²³I]Ioflupane scans without corresponding MRI, the analysis pipeline developed aimed to maximise the quality of spatial normalisation of the images in scenarios where only SPECT data was available.

A set of 17 SPECT templates, representing different stages of disease progression in PD, were generated in MNI152 space (Montreal Neurological Institute) to enable accurate normalisation of the [¹²³I]Ioflupane SPECT data. The 17 SPECT templates were derived from a healthy control SPECT template, with a range of progressively reduced signal intensities in the Caudate and Putamen, corresponding to increased deficits observed as PD progresses.

The healthy control SPECT template was constructed as follows; a customized brain atlas (containing 23 ROIs) specific to the dopamine transporter was firstly constructed in MNI152 space based on a sub-sample of the CIC brain atlas (including left and right subregions of the basal ganglia, including the Globus Pallidus, Ventral Pallidum, Pre-Caudate, Post-Caudate, Accumbens, Pre-dorsal Putamen, Pre-ventral Putamen, Post-dorsal Putamen, and Post-ventral Putamen, along with grey matter, white matter, cerebrospinal fluid (CSF), skull, and soft tissue ^23^. An optimisation algorithm then determined the optimal atlas region values and 3D Gaussian smoothing kernel required to best match to a healthy [¹²³I]Ioflupane SPECT image template in MNI152 space leading to the estimate of 23 regional values and a smoothing kernel of 12 mm full width at half maximum (FWHM). Subsequently, each voxel within the 23-region brain atlas was populated with the associated regional estimates and convolved with the 12mm FWHM smoothing kernel to generate the primary healthy control SPECT template. This process was then repeated using the parameters given in Table 1. to generate the remaining 16 disease specific templates.

**Table 1.**
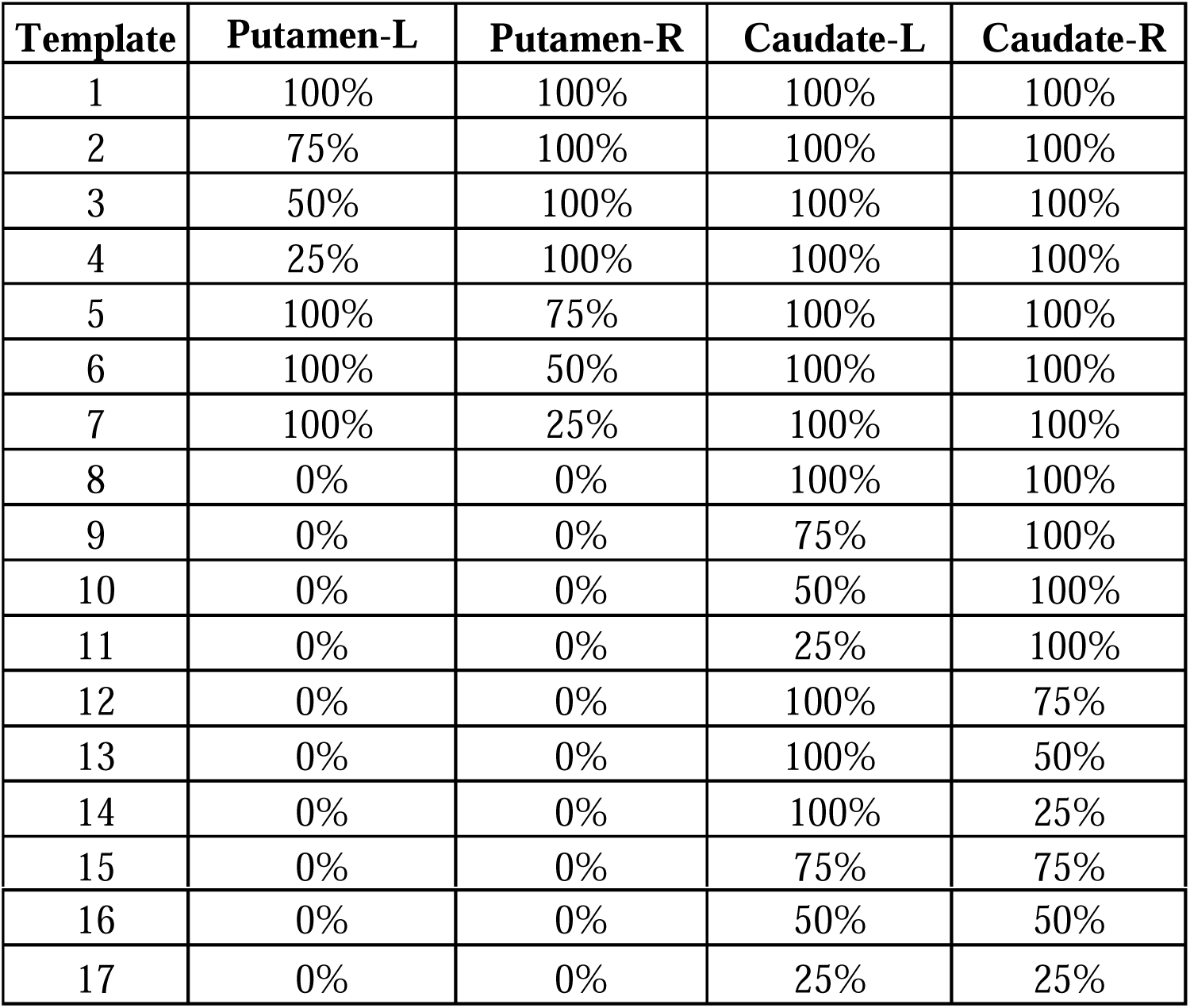
Definition of the 17 [¹²³I]Ioflupane SPECT templates as a percentage of the healthy control (HC) values in the Putamen and Caudate. All other regions contained 100% of the HC values.

Each [¹²³I]Ioflupane SPECT scan was spatially normalised into MNI152 space using an affine transformation to a linear combination (estimated as part of the normalisation process) of the 17 SPECT templates. A weighted image was also applied as part of this process to prioritize signals from the basal ganglia region, thereby enhancing the alignment of the SPECT data in relation to this critical area of interest. QC was performed on all images to ensure the quality of spatial normalisation.

### [^18^F]AV133 PET Analysis Pipeline

T1-MRI images were converted to 1mm isotropic format and brain segmentation into grey matter, white matter, and cerebrospinal fluid (CSF) was performed. DARTEL spatial normalization was then applied to derive mappings from the native space into MNI152 space using the standard T1 MNI152 template ^24^.

For the [^18^F]AV133 PET images, motion correction was applied using a frame-to-frame rigid registration algorithm, with the first frame set as the default reference image. Following realignment, an average image was then generated. The average [^18^F]AV133 image was then rigidly registered to the T1-MRI in native space and then spatially normalised into MNI152 space using the spatial transformation matrix derived previously. QC was performed on all images to ensure the quality of spatial normalisation.

### Calculation of SBR Outcome Measures

The reference (REF) and target (TAR) regions of interest from the CIC atlas were applied to the spatially normalized [¹²³I]Ioflupane SPECT and [^18^F]AV133 PET data in MNI152 space to derive the regional activity estimates SUV^REF^ and SUV^TAR^. This enabled quantification of regional SBR values for the chosen target regions (Left and Right) using the white matter as the reference region.

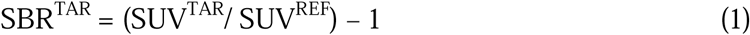

### The Centamine Scale

The Centamines approach consists of three levels. The first level outlines the methodology used to define the Centamine (CM) scale using [¹²³I]Ioflupane SPECT data, ranging from a baseline of 100% representing healthy controls to 0% corresponding to an absence of specific binding. The second and third level analyses then detail how other tracers may be converted into the Centamine scale using Head-to-Head data sets with either [¹²³I]Ioflupane SPECT data or a different tracer that has been previously calibrated to the Centamine scale using a Level 2 analysis.

### Level 1: Specification of Centamines with [¹²³I]Ioflupane SPECT data

The Level 1 analysis defines the Centamine scale based on healthy [¹²³I]Ioflupane SPECT data.

### Definition of Centamines

The Centamine scale provides a distinct Centamine value (CM) for a particular target region of interest (ROI). To enable this, it is necessary to define the mean SBR value from the cohort of healthy controls (n = 227) for this ROI (µ(*HC SBR*_*ROI*_^[¹²³I]*Ioflupane*^)). The Centamine value of this ROI for an individual [¹²³I]Ioflupane scan (with an SBR value in that region of *SBR*_*ROI*_^[¹²³I]*Ioflupane*^) can then be calculated as;

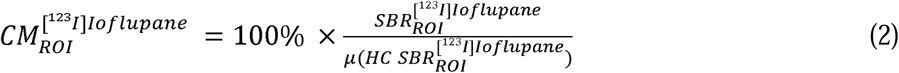

### Level 2: Mapping Tracer X into Centamines: Tracer X vs [¹²³I]Ioflupane SPECT

The Level 2 analysis enables mapping of a different dopaminergic neuronal imaging marker, Tracer X, into Centamines using a Head-to-Head data set acquired with [¹²³I]Ioflupane SPECT and Tracer X in the same subjects. It is envisaged that such a data set would include approximately 50 subjects with a suitable range of dopamine neuron levels to allow accurate estimation of the mapping. This number aligns with the prior Centiloids work ^15^.

A linear regression model is applied to the SBR Head-to-Head data from *Tracer X* and [¹²³*I*]*Ioflupane* scans for a given ROI, to derive the slope *a_Tracer X_* and intercept *b_Tracer X_* This relationship is given by,

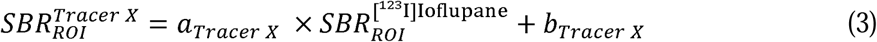

The *Tracer X SBR* values for CM=0% and CM=100% can then be determined for the given ROI by,

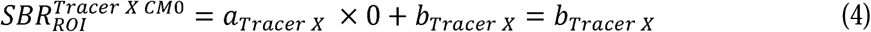

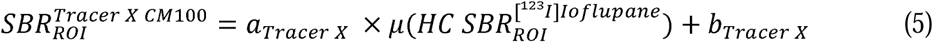

The Centamine value for an individual *Tracer X* scan can then be calculated as,

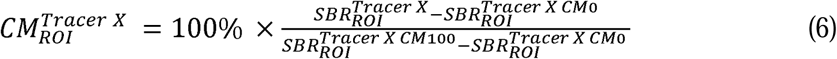

where *SBR_ROI_^Tracer X^* denotes *SBR* values for the individual *Tracer X* scan. By substituting the values for *SBR_ROI_^Tracer X CM0^* and *SBR_ROI_^Tracer X CM100^* from equations (4) and (5), the final form of the Centamine equation for *Tracer X* is,

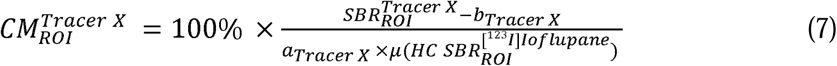

### Mapping [^18^F]AV133 PET into Centamines

Using this framework, a Level 2 analysis was performed with the [^18^F]AV133 PET vs [¹²³I]Ioflupane SPECT Head-to-Head data set involving 68 subjects with a total of 162 paired scans (Data Set 2). In this analysis, all the paired available scans were treated as independent data points.

### Level 3: Mapping Tracer Y into Centamines: Tracer Y vs Tracer X

The Level 3 analysis enables mapping of a different dopaminergic neuronal imaging marker, *Tracer Y*, into Centamines using a Head-to-Head data set acquired with *Tracer Y* and *Tracer X* in the same subjects. This is conditional on *Tracer X* having previously undergone a Level 2 analysis using a *Tracer X* vs [¹²³I]Ioflupane SPECT Head-to-Head data set. Again, it is envisaged that such a data set would include approximately 50 subjects with a suitable range of dopamine neuron levels to allow accurate estimation of the mapping. A linear regression model is applied to the SBR Head-to-Head data from *Tracer Y* and *Tracer X* scans for a given ROI, to derive the slope *a_Tracer Y_* and intercept *b_Tracer v_*. This relationship is given by,

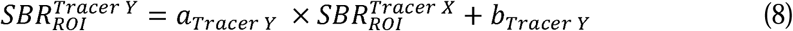

Using known *Tracer X* values for CM=0% and CM=100% derived from the Level 2 analysis, the corresponding *Tracer Y SBR* values for CM=0% and CM=100% can be determined for the given ROI as,

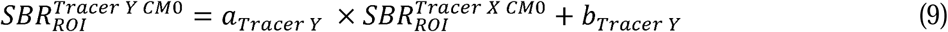

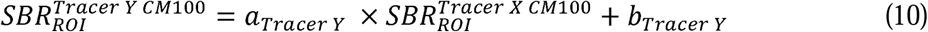

The Centamine value for an individual *Tracer Y* scan is then be calculated by,

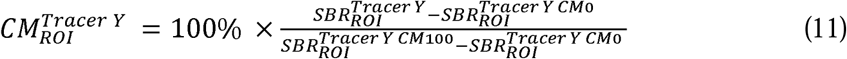

where *SBR_ROI_^Tracer r^* denotes SBR values for the individual *Tracer Y* scan. By substituting the values for *SBR_ROI_^Tracer Y CM0^*, *SBR_ROI_^TracerY CM100^*, *SBR_ROI_^Tracer X CM0^* and *SBR_ROI_^Tracer X CM100^* from Equations (4), (5), (9) and (10), the final form of the Centamine equation for *Tracer Y* is

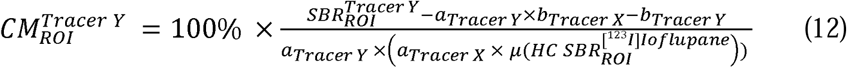

### Correction for Age and Sex Effects

Prior harmonisation approaches, such as Centiloids ^15^ and Centaurs ^19^, have not corrected their outcome measures for age and sex effects and we align with those approaches here in terms of the definition of CM. However, there may be applications like stratification or diagnostics, using for instance the Putamen, that want to explicitly correct for these effects given that dopaminergic neuronal density as measured by DAT has demonstrated a modest reduction over time (approx. 5% per decade) ^17,25,26^; and sex differences (females approx. 10-15% higher) ^26^. To this end, we propose an example of age and sex corrected Centamine (CM^*^) outcome measure that may be further helpful in these applications. This transformation is determined from the Level 1 output data with DAT in Centamines from healthy controls. A linear regression is performed on these data using the following equation,

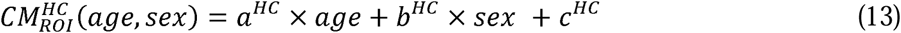

where ***CM_ROI_*** is the Centamine value in ROI, age is the age at the time of the scan and sex is 0 for female and 1 for male subjects. We propose to anchor ***CM***^*^ to the expected average ***CM*** value for a 65-year-old person which yields the following equation,

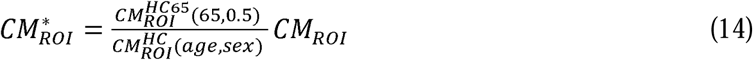

or explicitly,

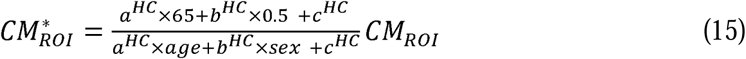

where age and sex are the actual age and sex of the subject whose Centamine value is CM_ROI_.

## RESULTS

All [¹²³I]Ioflupane SPECT and [^18^F]AV133 scans were successfully spatially normalised into MNI152 space, as assessed by visual assessment, and SBR values were calculated for left and right Striatum, Putamen, Caudate, Pre-Commissural Putamen and Post-Commissural Putamen.

### Level 1 Analysis: [¹²³I]Ioflupane SPECT

The results from the Level 1 analysis of the [¹²³I]Ioflupane SPECT data to derive the SBR anchor points for Centamine values of 0 and 100% for [¹²³I]Ioflupane SPECT are provided in Table 2.

**Table 2.**
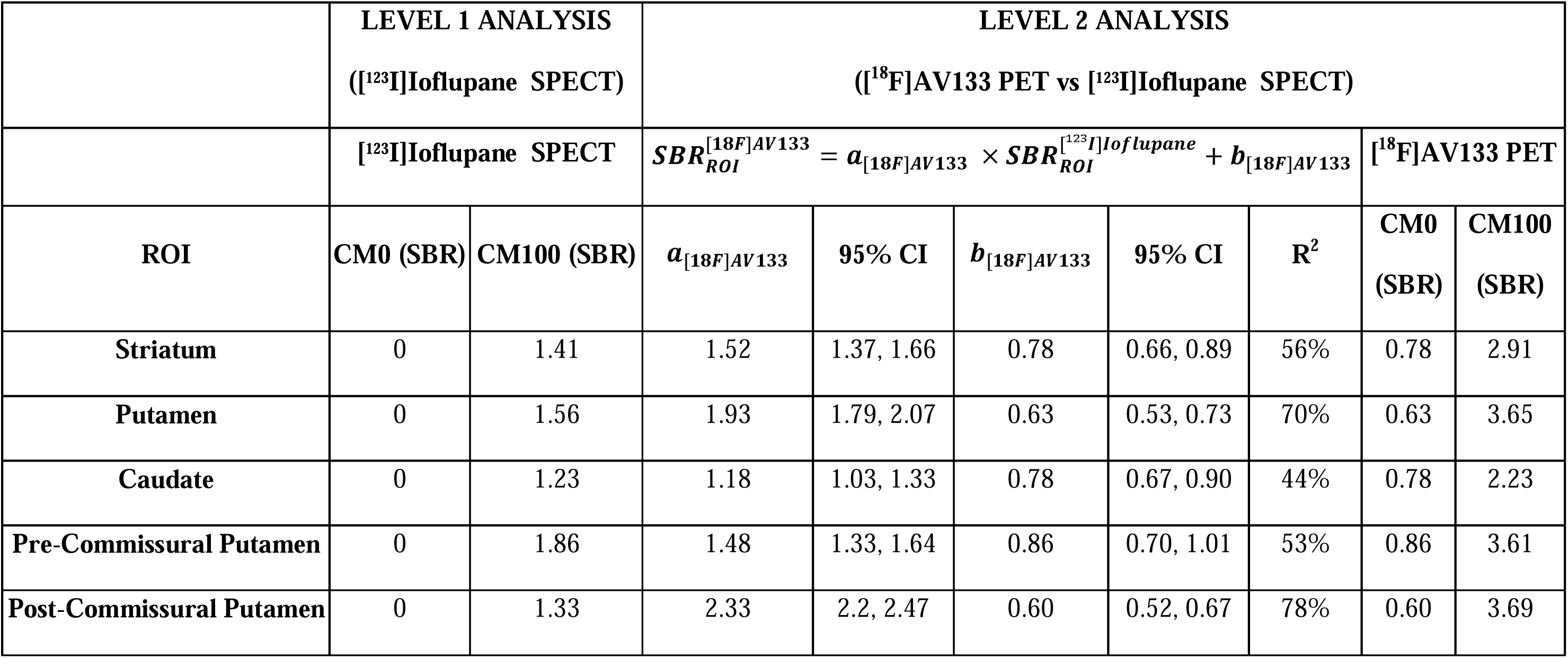
Results of the Level 1 and Level 2 analyses detailing the specific binding ratio (SBR) anchor points of CM=0% (CM0) and CM=100% (CM100) for [¹²³I]Ioflupane SPECT and [^18^F]AV133 PET for the five target regions of interest.

### Level 2 Analysis: [^18^F]AV133 PET vs [¹²³I]Ioflupane SPECT

The results from the Level 2 analysis of the [^18^F]AV133 PET and [¹²³I]Ioflupane SPECT to derive the SBR anchor points for Centamine values of 0 and 100% for [^18^F]AV133 PET are provided in Table 2. The linear regressions for the five target regions are displayed in Figure 3 along with their subsequent transformation into Centamines demonstrating a moderate-strong relationship between [^18^F]AV133 PET and [¹²³I]Ioflupane SPECT. The calculated regional R^2^ ranged from 44-78%, with the highest value observed in the Post-Commissural Putamen and the lowest value in the Caudate. The highest slope and lowest intercept were both obtained in the Post-Commissural Putamen. Overall, the most important regions for PD research applications (Post-Commissural Putamen, Putamen and Striatum) provided the best agreements. Additional information on the analysis of the agreement between the Centamine values are provided in Supplementary Figures 1-3 which include Bland–Altman plots, histograms of the Centamine values and the individual differences across [^18^F]AV133 PET and [¹²³I]Ioflupane SPECT.

**Figure 3.**
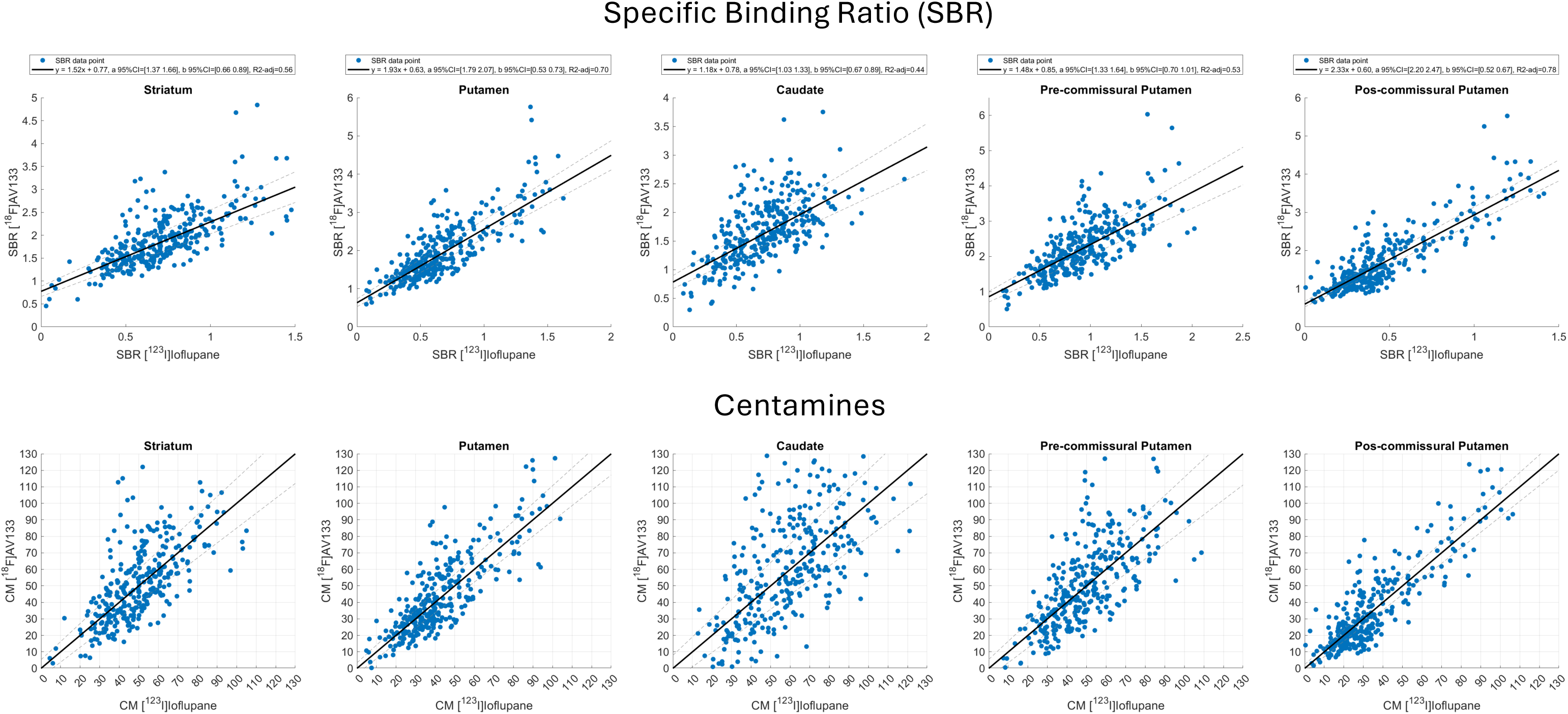
Results from the Level 2 regression analyses of the Head-to-Head [^18^F]AV133 PET and [¹²³I]Ioflupane SPECT SBR data (top row) and transformation of the data into the Centamine scale (bottom row) for the five target regions of interest. Note that a small number of points are outside the axis range of 0-130 in the Centamine plots and are not displayed.

Figure 4 illustrates all of the data mapped into Centamines and confirms a good alignment of the Head-to-Head [^18^F]AV133 PET and [¹²³I]Ioflupane SPECT data across all five regions with a minimum group-level difference in Post-Commissural Putamen (1.5%) and a maximum group difference in Caudate (3%). The Head-to-Head data demonstrated an increasing reduction in Centamines from Caudate (lowest reduction), through Striatum, Pre-Commissural Putamen and Putamen, to Post-Commissural Putamen (highest reduction) consistent with the prevalence of PD subjects in this cohort and the known loss of dopaminergic neurons due to the disease.

**Figure 4.**
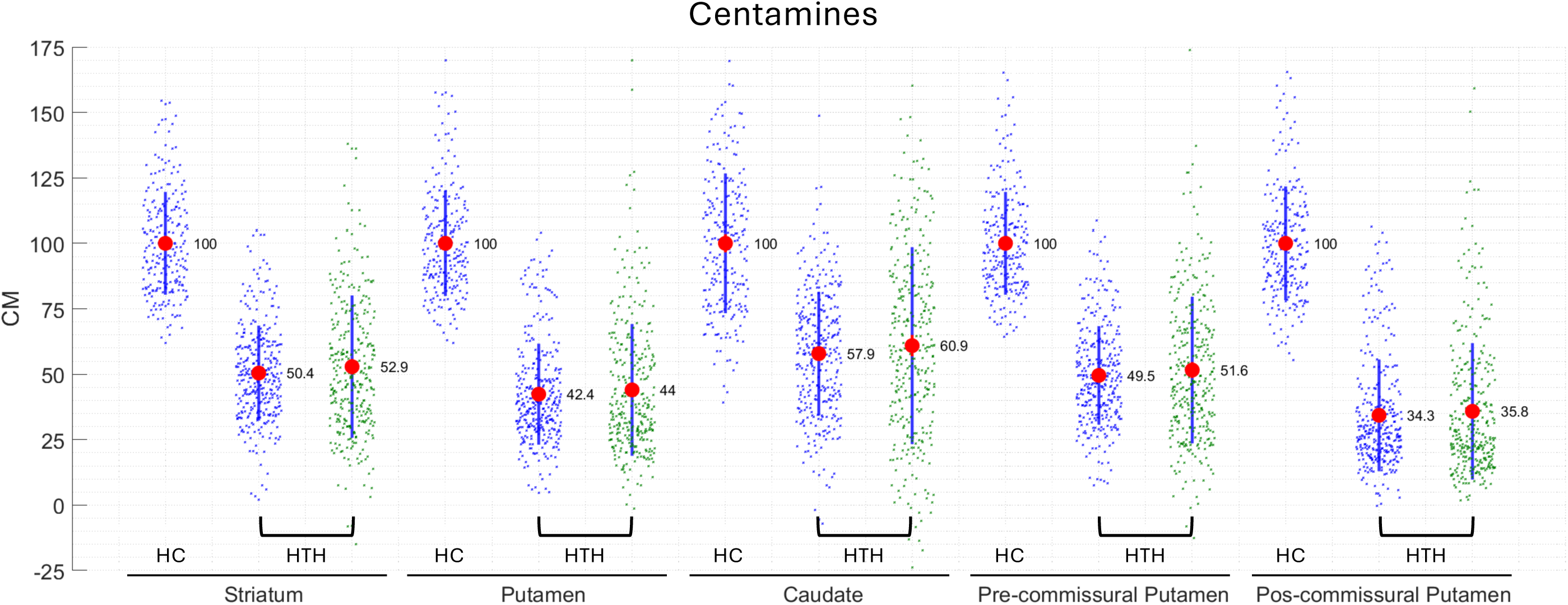
All data from Data Sets 1 & 2 mapped into the Centamine scale following Level 1 & 2 analyses (Data Set 1 (HC), Data Set 2 (HTH), [¹²³I]Ioflupane SPECT (Blue), [^18^F]AV133 PET (Green), group means displayed as red dots and black lines indicate +/- SD).

As there was longitudinal data present in the Head-to-Head data set (94 follow up scans were included), it was also possible to perform a longitudinal analysis to calculate the mean change per annum, using linear regression analysis, for each imaging marker following Centamine transformation. The results for the mean change are given in Table 3 and demonstrate that this was similar for both imaging markers.

**Table 3.**
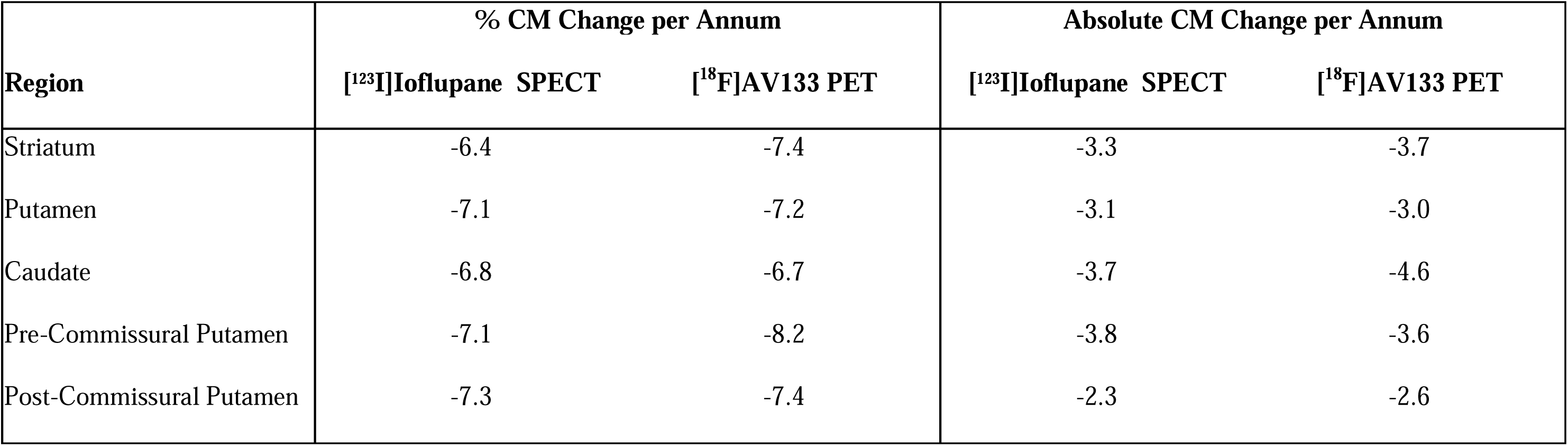
Longitudinal analysis of the Head-to-Head data, transformed into Centamines, to calculate mean changes per annum.

### Correction for Age and Sex Effects

Linear regression analyses were performed on the Centamine values derived from the healthy [¹²³I]Ioflupane SPECT data (n=227) for Putamen regions with age and sex as regressors using equation 13 (Figure 5.). This then enabled calculation of the age and sex corrected Centamine values (CM^*^) using equation 15. The Putamen demonstrated a 14.8% higher Centamine values in females and a reduction of approximately 2.4% per decade in both sexes. Similar results were obtained for the subregions of the Putamen with slightly higher age-related decline in the Pre-Commissural Putamen (2.7%/decade) as compared to the Post-Commissural Putamen (2.1%/decade). The comparison of the group distributions for CM^*^ vs CM can be seen in Figure 5 where it is evident that there is a modest reduction in variance and alignment of means with 100% for both male and females. Whilst modest, this correction will likely be important for stratification/diagnostic applications employing a cut-off value to determine a classification of dopamine neuron deficit.

**Figure 5.**
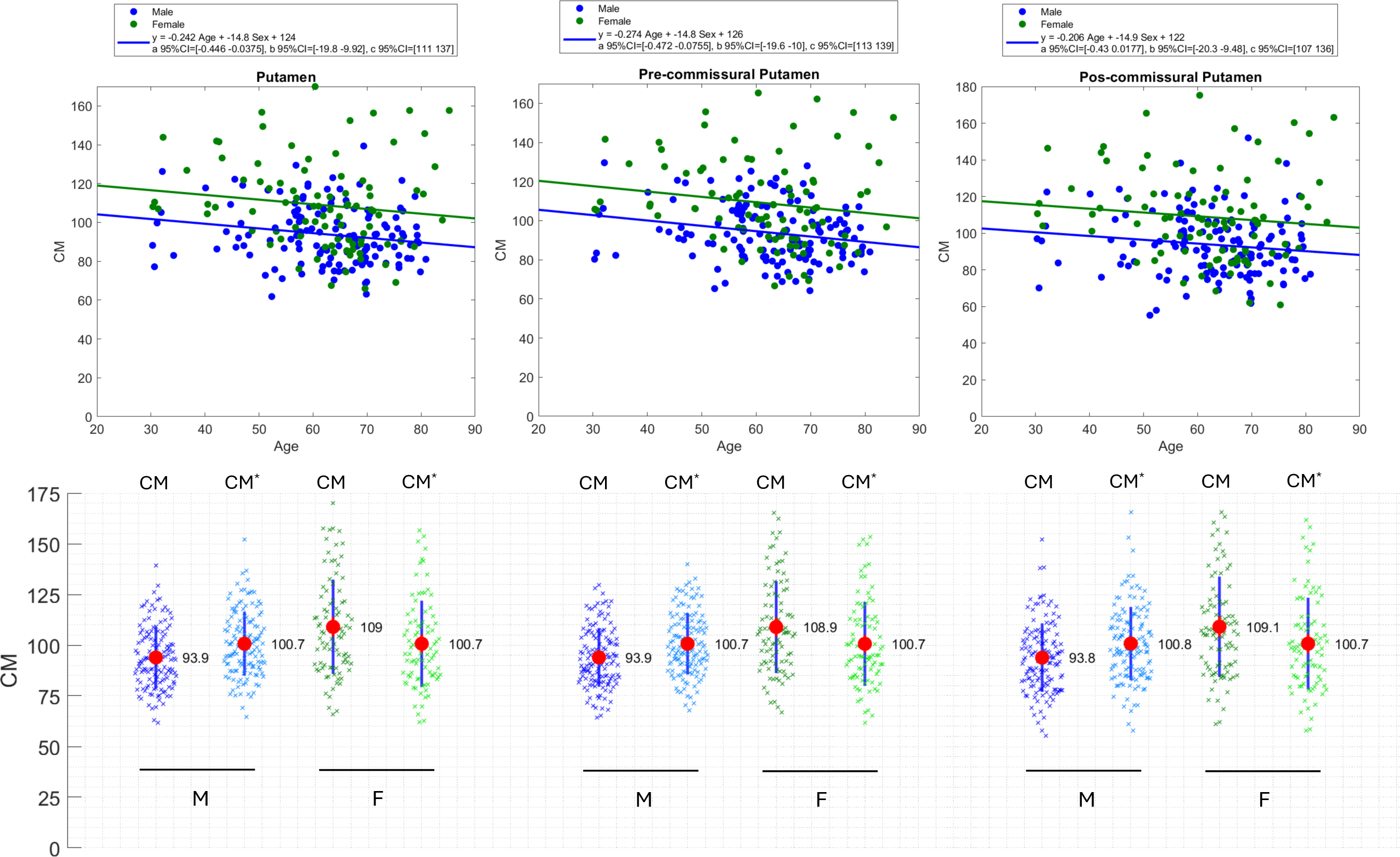
Linear regression analysis of the healthy [¹²³I]Ioflupane SPECT data with age and sex as regressors for Putamen regions (top) and. Centamine (CM) and age/sex corrected Centamine (CM*) values for male (M) and female (F) groups (bottom).

## DISCUSSION

The Centamine scale has been proposed to enable a harmonized quantification framework for dopaminergic imaging with molecular imaging tracers. The Centamine methodology will enable multiple dopamine neuron imaging biomarkers to be compared and even for multiple imaging biomarkers to be used in a single clinical study potentially informing and accelerating PD clinical trials throughout the course of disease. This approach builds on the success of Centiloids and Centaurs, which have proposed and developed harmonized amyloid- and tau-PET quantification in AD, simplifying eligibility assessment, enhancing longitudinal data reporting and improving cross-tracer comparability.

Developing the Centamine scale is particularly timely given the recent paradigm shift to a biologic definition of PD and related synucleinopathies and the integrated biologic and clinical NSD-ISS^2^. Dopamine imaging is a key anchor biomarker for NSD staging.

Understanding the earliest stages of dopamine dysfunction may enable studies investigating molecular determinants of neurodegeneration as disease progresses. The availability of a standardized quantitative dopamine imaging outcome would provide an intuitive scale to enable clear and reliable disease staging for research studies. The Centamine strategy would also accelerate therapeutic studies by enabling multiple tracers to be used in a single study. Centamines would further enable comparison of dopaminergic neuronal loss in different brain regions that might inform disease progression throughout the disease both prior to typical symptoms and after functional impairment is present. Finally, Centamines would also allow an easier comparison between clinical therapeutic trials and potentially between different clinical syndromes, for example PD and MSA.

The Centamine approach follows a similar principle to Centiloids and Centaurs, but with key differences. Unlike amyloid and tau, which accumulate over time with disease progression, dopaminergic neuron levels decline from a normal baseline. As a result, the Centamine scale is inverted, with 100% representing a healthy state, whereas in Centiloids and Centaurs, 100% corresponds to the average amyloid- and tau-PET signal in typical AD. This inversion somewhat simplifies the definition of the Centamine scale, as it can be directly established from a healthy population without needing to anchor 100% to a specific disease stage, as required by Centiloids and Centaurs. It should be noted that the CM* scale for [¹²³I]Ioflupane SPECT is related to the percent deviation reported by DaTQUANT^TM^ software ^27,28^, which instead anchors the mean of (age-dependent) normal to be 0 and the absence of signal (SBR=0) to be 1.

Furthermore, Centamine values are by necessity region-specific, given the heterogeneous and region-specific loss of dopamine neuron levels in PD. This allows for multiple Centamine values per scan, making it functionally closer to Centaurs, which also accounts for regional variation, rather than Centiloids, which assigns a single value per scan due to the uniform accumulation rate of amyloid across the brain ^29^. This offers another opportunity to use Centamines to investigate differences in regional pathology in PD and related synucleinopathies.

The Centamine framework consists of three levels of analysis to standardize the quantification of dopaminergic imaging tracers. This study presents a Level 1 analysis, which defines the Centamine scale using a large cohort of healthy individuals (n=227) imaged with [¹²³I]Ioflupane SPECT. A Level 2 analysis was conducted using Head-to-Head imaging with [^18^F]AV133 PET and [¹²³I]Ioflupane SPECT, enabling the additional conversion of [^18^F]AV133 PET into the Centamine scale. The framework also outlines a Level 3 analysis, which facilitates the conversion of additional tracers without requiring a direct Head-to-Head [¹²³I]Ioflupane SPECT dataset. Instead, it leverages a Head-to-Head comparison with another tracer that has already undergone a Level 2 analysis, ensuring continuity in tracer standardization.

The framework enables the harmonization of additional dopaminergic imaging markers, with ongoing Head-to-Head studies aimed at further expanding the Centamine scale. Current studies include [^18^F]PE2I PET vs [¹²³I]Ioflupane SPECT ([^18^F]PE2I PET through a Level 2 analysis) and [^18^F]FPCIT PET vs [^18^F]AV133 PET ([^18^F]FPCIT through a Level 3 analysis). These datasets will support the future integration of these tracers into the Centamine scale, increase our understanding of individual tracer signal-to-noise and expand its applicability.

As with many studies, there are certain limitations. In theory, if two molecular imaging markers measure the same target in the same person, their specific binding results (SBR) may be expected to follow a simple mathematical relationship: *y = ax*. However, in this study, an intercept term was needed to accurately describe the relationship between [¹[F]AV133 PET and [¹²³I]Ioflupane SPECT. This intercept was fairly consistent across the five brain regions analyzed (0.60–0.86, see Table 1). It may be due to factors like differences in non-specific binding between target and reference regions, limitations in [¹²³I]Ioflupane SPECT’s sensitivity at low dopamine levels, or variations in DAT and VMAT2 expression as the disease progresses. The need for an intercept is not surprising, as similar findings have been seen in studies using Centiloids and Centaurs. The future analysis of the [^18^F]PE2I PET vs [¹²³I]Ioflupane SPECT and [^18^F]FPCIT PET vs [^18^F]AV133 PET data sets will provide further insights into these relationships. In summary, this study confirms a significant relationship between [¹[F]AV133 PET and [¹²³I]Ioflupane SPECT, with the Centamine scale providing a useful way to align them.

The proposed approach is designed to allow individual research groups to calibrate their own image analysis pipelines, ensuring they can generate results in Centamines. To enable this, the requisite healthy [¹²³I]Ioflupane SPECT and Head-to-Head data sets will be made available (see http://ppmi-info.org for details).

We have successfully developed the Centamine scale for harmonized quantification of dopaminergic neuronal imaging markers in Parkinson’s Disease and related synucleinopathies with the objective of further enabling and accelerating clinical therapeutic trials and diagnostic applications.

## Supporting information

PPMI Study Group Authors

## Data Availability

Data used in the preparation of this article were obtained on [2025-03-28] from the Parkinson's Progression Markers Initiative (PPMI) database (https://www.ppmi-info.org/access-data-specimens/download-data), RRID:SCR_006431. For up-to-date information on the study, visit http://www.ppmi-info.org.

## AUTHOR CONTRIBUTIONS

1. Conception and design of the study
2. Acquisition and analysis of data
3. Drafting a significant portion of the manuscript or figures

Zhen Fan 1, 2, 3; Graham Searle 2, 3; Gaia Rizzo 2, 3; Justin Albani 2, 3; Patrick Cella 3; Robert Comley 3; Gregory Klein 3; Luca Passamonti 3; Cristian Salinas 3; Adam J. Schwarz 3; Leonardo Iaccarino 3; Gilles Tamagnan 3; Jamie Eberling 1, 3; Ken Marek 1, 3; John Seibyl 1, 3; Roger N. Gunn 1, 2, 3.

## Acknowledgement statement (including conflict of interest and funding sources)

This work was funded by The Michael J. Fox Foundation for Parkinson’s Research (Grant ID: MJFF-025525). Data used in the preparation of this article were obtained on [2025-03-28] from the Parkinson’s Progression Markers Initiative (PPMI) database (https://www.ppmi-info.org/access-data-specimens/download-data), RRID:SCR_006431. For up-to-date information on the study, visit http://www.ppmi-info.org. We acknowledge that PPMI is scientifically supported and largely funded by The Michael J Fox Foundation for Parkinson’s Research and Aligning Science Across Parkinson’s and the PPMI Partners Scientific Advisory Board (https://www.ppmi-info.org/about-ppmi/who-we-are/study-sponsors).

PPMI – a public-private partnership – is funded by the Michael J. Fox Foundation for Parkinson’s Research and funding partners, including 4D Pharma, Abbvie, AcureX, Allergan, Amathus Therapeutics, Aligning Science Across Parkinson’s, AskBio, Avid Radiopharmaceuticals, BIAL, BioArctic, Biogen, Biohaven, BioLegend, BlueRock Therapeutics, Bristol-Myers Squibb, Calico Labs, Capsida Biotherapeutics, Celgene, Cerevel Therapeutics, Coave Therapeutics, DaCapo Brainscience, Denali, Edmond J. Safra Foundation, Eli Lilly, Gain Therapeutics, GE HealthCare, Genentech, GSK, Golub Capital, Handl Therapeutics, Insitro, Jazz Pharmaceuticals, Johnson & Johnson Innovative Medicine, Lundbeck, Merck, Meso Scale Discovery, Mission Therapeutics, Neurocrine Biosciences, Neuron23, Neuropore, Pfizer, Piramal, Prevail Therapeutics, Roche, Sanofi, Servier, Sun Pharma Advanced Research Company, Takeda, Teva, UCB, Vanqua Bio, Verily, Voyager Therapeutics, the Weston Family Foundation and Yumanity Therapeutics.

## REFERENCES

1. Siderowf A, Concha-Marambio L, Lafontant DE, et al. Assessment of heterogeneity among participants in the Parkinson’s Progression Markers Initiative cohort using α-synuclein seed amplification: a cross-sectional study. Lancet Neurol 2023;22(5):407– 417.

2. Simuni T, Chahine LM, Poston K, et al. A biological definition of neuronal α-synuclein disease: towards an integrated staging system for research. Lancet Neurol 2024;23(2):178–190.

3. Höglinger GU, Adler CH, Berg D, et al. A biological classification of Parkinson’s disease: the SynNeurGe research diagnostic criteria. Lancet Neurol 2024;23(2):191– 204.

4. Booij J, Tissingh G, Boer GJ, et al. [123I]FP-CIT SPECT shows a pronounced decline of striatal dopamine transporter labelling in early and advanced Parkinson’s disease. J Neurol Neurosurg Psychiatry 1997;62(2):133–140.

5. Darcourt J, Booij J, Tatsch K, et al. EANM procedure guidelines for brain neurotransmission SPECT using (123)I-labelled dopamine transporter ligands, version 2. Eur J Nucl Med Mol Imaging 2010;37(2):443–450.

6. Benamer HTS, Patterson J, Grosset DG, et al. Accurate differentiation of parkinsonism and essential tremor using visual assessment of [123 I]-FP-CIT SPECT imaging: The [123 I]-FP-CIT study group. Mov Disord 2000;15(3):503–510.

7. Kong Y, Zhang C, Liu K, et al. Imaging of dopamine transporters in Parkinson disease: a meta-analysis of 18F/123I-FP-CIT studies. Ann Clin Transl Neurol 2020;7(9):1524–1534.

8. Wang J, Zuo CT, Jiang YP, et al. 18F-FP-CIT PET imaging and SPM analysis of dopamine transporters in Parkinson’s disease in various Hoehn & Yahr stages. J Neurol 2007;254(2):185–190.

9. Sasaki T, Ito H, Kimura Y, et al. Quantification of Dopamine Transporter in Human Brain Using PET with 18F-FE-PE2I. Journal of Nuclear Medicine 2012;53(7):1065– 1073.

10. Delva A, Van Weehaeghe D, van Aalst J, et al. Quantification and discriminative power of 18F-FE-PE2I PET in patients with Parkinson’s disease. Eur J Nucl Med Mol Imaging 2020;47(8):1913–1926.

11. Alexander PK, Lie Y, Jones G, et al. Management Impact of Imaging Brain Vesicular Monoamine Transporter Type 2 in Clinically Uncertain Parkinsonian Syndrome with 18F-AV133 and PET. Journal of Nuclear Medicine 2017;58(11):1815–1820.

12. Beauchamp LC, Dore V, Villemagne VL, et al. Using 18F-AV-133 VMAT2 PET Imaging to Monitor Progressive Nigrostriatal Degeneration in Parkinson Disease. Neurology 2023;101(22):E2314–E2324.

13. Okamura N, Villemagne VL, Drago J, et al. In Vivo Measurement of Vesicular Monoamine Transporter Type 2 Density in Parkinson Disease with 18F-AV-133. Journal of Nuclear Medicine 2010;51(2):223–228.

14. Fan Z, Searle G, Whittington A, et al. Increasing the power of [18F]AV133 PET to measure longitudinal changes in Parkinson’s Disease: Identification of optimal reference and target regions. AD/PD^TM^ International Conference on Alzheimer’s and Parkinson’s Diseases and related neurological disorders, Gothenburg, Sweden. 2023;

15. Klunk WE, Koeppe RA, Price JC, et al. The Centiloid Project: Standardizing Quantitative Amyloid Plaque Estimation by PET. Alzheimers Dement 2014;11(1):1.

16. Collij LE, Bollack A, La Joie R, et al. Centiloid recommendations for clinical context-of-use from the AMYPAD consortium. Alzheimers Dement 2024;20(12)

17. van Dyck CH. Age-Related Decline in Dopamine Transporters: Analysis of Striatal Subregions, Nonlinear Effects, and Hemispheric Asymmetries. American Journal of Geriatric Psychiatry 2002;10(1):36–43.

18. Sims JR, Zimmer JA, Evans CD, et al. Donanemab in Early Symptomatic Alzheimer Disease: The TRAILBLAZER-ALZ 2 Randomized Clinical Trial. JAMA 2023;330(6):512–527.

19. Leuzy A, Raket LL, Villemagne VL, et al. Harmonizing tau positron emission tomography in Alzheimer’s disease: The CenTauR scale and the joint propagation model. Alzheimer’s & Dementia 2024;20(9):5833–5848.

20. M Tanner C, Marek K. Parkinson’s Progression Markers Initiative Online Study (PPMI Online) v1. 2024;

21. Marek K. Early Longitudinal Imaging in the Parkinson’s Progression Markers Initiative Using [ 18F] AV-133 (PPMI AV-133 Prodromal Imaging) v1. 2024;

22. Gunn R, Coello C, Searle G. Molecular Imaging And Kinetic Analysis Toolbox (MIAKAT) - A Quantitative Software Package for the Analysis of PET Neuroimaging Data. Journal of Nuclear Medicine 2016;57(supplement 2):1928–1928.

23. Tziortzi AC, Searle GE, Tzimopoulou S, et al. Imaging dopamine receptors in humans with [11C]-(+)-PHNO: dissection of D3 signal and anatomy. Neuroimage 2011;54(1):264–277.

24. Ashburner J. A fast diffeomorphic image registration algorithm. Neuroimage 2007;38(1):95–113.

25. Van Dyck CH, Seibyl JP, Malison RT, et al. Age-Related Decline in Striatal Dopamine Transporter Binding with Iodine-123-β-CITSPECT. Journal of Nuclear Medicine 1995;36(7):1175–1181.

26. Lavalaye J, Booij J, Reneman L, et al. Effect of age and gender on dopamine transporter imaging with [123I]FP-CIT SPET in healthy volunteers. Eur J Nucl Med 2000;27(7):867–869.

27. Neill M, Fisher JM, Brand C, et al. Practical Application of DaTQUANT with Optimal Threshold for Diagnostic Accuracy of Dopamine Transporter SPECT. Tomography 2021;7(4):980–989.

28. Kuo PH, Cella P, Chou YH, et al. Optimal DaTQUANT Thresholds for Diagnostic Accuracy of Dementia with Lewy Bodies (DLB) and Parkinson’s Disease (PD). Tomography 2024;10(10):1608–1621.

29. Whittington A, Sharp DJ, Gunn RN. Spatiotemporal Distribution of β-Amyloid in Alzheimer Disease Is the Result of Heterogeneous Regional Carrying Capacities. Journal of Nuclear Medicine 2018;59(5):822–827.

