## Supplementary figures and images for "Towards Harmonizing Quantification of Dopamine Neuron Imaging Biomarkers in Parkinson’s Disease: The Centamine Scale"

### Supplement Materials

#### Supplementary Figure 1

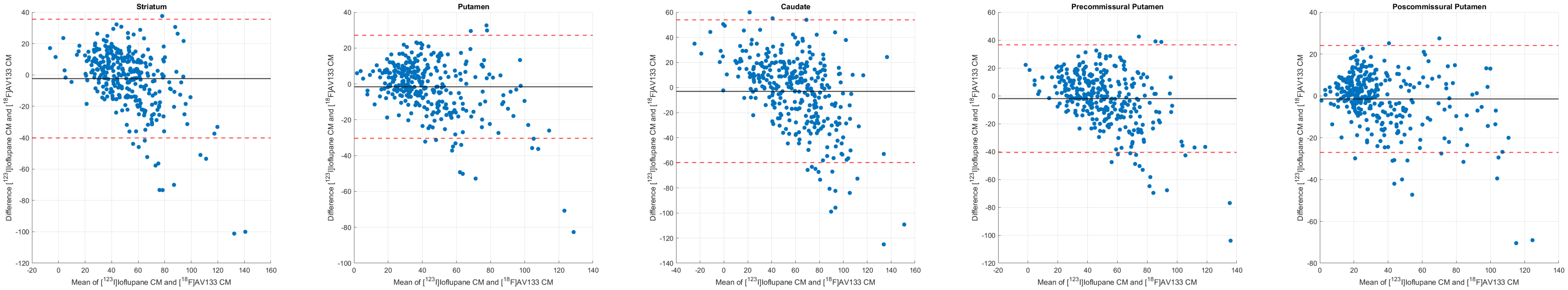

Supplementary Figure 2

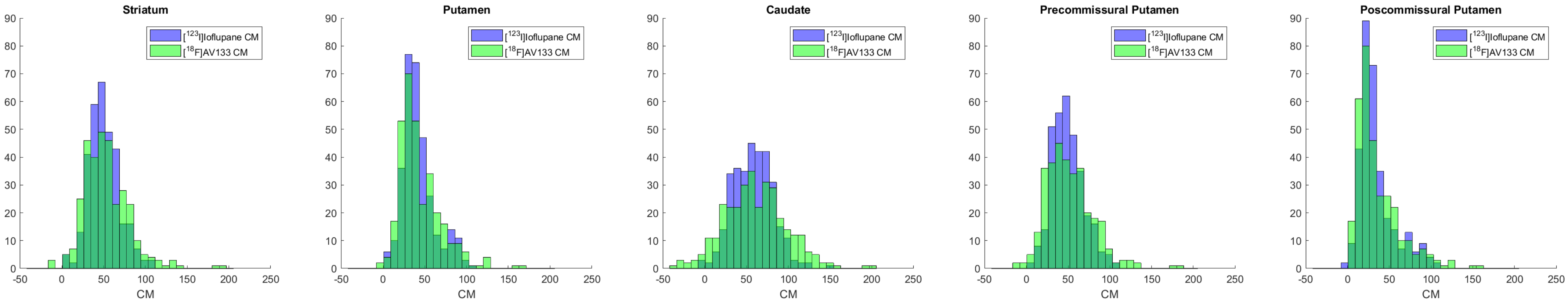

Supplementary Figure 3

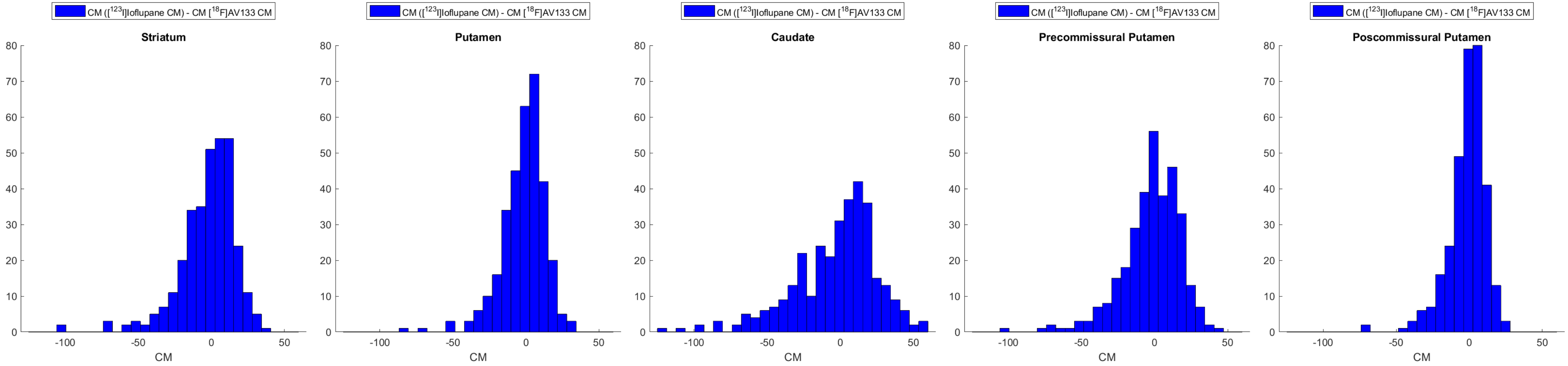
